# Assessment of the implementation of iron and folic acid tablets supplementation to in-school girls in Nanumba South District of Northern Region, Ghana

**DOI:** 10.1101/2024.11.26.24317998

**Authors:** Nayina Bakunboa Isaac Razak, Robert Kogi, Jean Claude Ndayishimiye, Samuel Yaw Opoku

**Author notes:** **Corresponding Author:** Nayina Bakunboa Isaac Razak, **Email:**.

## Abstract

Iron deficiency is a leading cause of morbidity and mortality among adolescent girls 10– 19 years of age globally. Despite the free weekly iron and folic acid supplementation, prevalence of anaemia among adolescent females still between 24% to 64.6%. This study sought to identify the factors affecting the implementation of iron and folic acid program to a school-based anaemia reduction in the Nanumba South District, Northern Region, Ghana.

A cross-sectional analytical study was carried out. And a simple random sampling method was used to select the schools and the adolescent girls, while purposive sampling was used to select school health coordinators and health staff. The outcome variable was IFA compliance and exposures were knowledge on anaemia and IFAS program, as well as socio-demographic factors (Age, marital status, level of education, gestational age, parity, gestation age, and occupation). Adolescent girls who consumed at least five tablets of the expected dose in the previous 7 weeks (1 tablet per week) which is equivalent to consuming 70% of the expected dose before the day of the data collection were considered compliant. Respondents who were able to get “1” in all the four knowledge questions were considered to have good level of knowledge. Occupation level of parents was self-identified by the respondents. Multivariable logistic regression analysis was performed to identify the predictors of the outcome variable. Significance level was set at p<0.05.

The level of knowledge about benefits of iron and folic acid supplementation was generally high in this study (88.0%). Overall, the majority (65.7%) of the respondents complied with iron and folic acid supplementation. Father’s occupational level was the only variable, which was significantly associated with adolescent girls’ IFA consumption compliance (p=0.002). The odds of compliance among respondents who were 17-25 years was significantly 56% times lower compared to those who were 10-16 years (AOR = 0.44 (95% CI: 0.21-0.96) p = 0.038). Adolescent girls of Kokomba ethnic group were also 83% less likely to comply with iron and folic acid consumption (AOR = 0.17 (95% CI: 0.03-0.89) p = 0.036).

In conclusion, the study highlighted high compliance with the IFA among respondents, emphasizing the critical roles of teachers and health workers in counseling, monitoring, and supervision, while identifying age 17-25 years and Kokomba ethnic group being less significant factors influencing compliance. It is recommended that the role of teachers and health workers in the IFAS program should be strengthened, while addressing barriers to compliance among older adolescents and engaging families.

## Introduction

Iron deficiency anaemia in adolescents remains a major public health challenge. The prevalence of nutritional anaemia among adolescent girls in developing countries, particularly in low- and middle-income countries (LMICs), is alarmingly high. In West and Central Africa, the prevalence of adolescent anaemia remained high at 45% over the last two decades [1]. A broader analysis across twenty-four (24) LMICs indicated anaemia pooled prevalence of 41.58% among young women aged 15-24 [2].

While adolescence is a time of remarkable growth and development, iron deficiency can impair growth and delay sexual menstruation among young girls [3]. During this time, about 20% of final adult height and 50% of adult weight are attained [4]. Due to their increased nutritional requirements during growth and development as well as menstrual blood loss, adolescent girls are more likely to develop iron deficiency related anemia [5]. To prevent anaemia and maintain growth during adolescence, the weekly intermittent iron and folic acid supplementation (IFA) has been recommended by the World Health Organization [6]. It entails providing adolescent girls a supplement once each week for a specific amount of time that contains both iron and folic acid [7].

Despite the free weekly iron and folic acid supplementation, prevalence of anaemia among adolescent females remains between 24% [8] to 64.6% [9]. Anaemia has complex and long-term consequences, especially in adolescent girls, and it is also related to children morbidity and mortality [10]. To encourage the intake of IFA tablets across many settings and populations, these are provided free of charge utilizing school institutions as a distribution channel for adolescent girls as one of the target groups. In Ghana, the Girls’ Iron-Folic acid Tablet Supplementation (GIFTS) program, which is an integrated health and nutrition education program with intermittent weekly IFA supplementation, was designed to reach adolescent girls ages 10–19 years simultaneously through two delivery platforms: schools and local health centers. In October 2017, the first phase of the program began in the Brong-Ahafo, Northern, Upper East, and Volta regions of Ghana [11].

The success of the iron supplementation program depends largely on the effectiveness of a delivery system and the compliance of the target recipients. The school is the most efficient channel for the delivery of health services to schoolchildren and is considered a high-priority area for public health education [12, 13,14]. However, the effectiveness of programs that aim to supplement iron has been questioned because of the low efficiency of health services and poor compliance of the target recipients [15]. Undeniably, there have been several studies of the barriers and facilitators to compliance with IFA supplementation in the GIFTS program in many settings in Ghana [16,17,18]. Unfortunately, none of these studies has been conducted among adolescent girls in the Nanumba South District. Hence, this study was carried out to assess the factors affecting the implementation of IFA program to a school-based anaemia reduction program with weekly iron and folic acid supplementation in the Nanumba South District.

## Methods

### Study Design

A school-based cross-sectional analytical study was used in carrying out this study. This design was chosen because we intended to gain immediate knowledge and information on IFAS in the Nanumba South District.

### Study Population

All Adolescent girls attending junior high school in the Nanumba South District were the source population. The study population consisted of randomly sampled girls who were attending junior high school in the study area in the third term of the 2022/2023 academic year. Junior high school students were used because they are typically adolescents, who are at a critical growth stage with increased nutritional demands [19]. In addition, adolescents, particularly girls, are more prone to iron deficiency anaemia due to factors such as menstruation and rapid physical growth [20,21,22].

Girls who were absent and those who declined to participate in the study were excluded. School health coordinators of the randomly selected schools and health staff were also selected to participate in the study.

### Sample Size Determination and Sampling Technique

The sample size was determined using Cochran’s formula N= (Z^2^ × p(1 − p))/d2 [23] with 95% confidence interval and 5% margin of error with compliance rate of 26.2% [16] as reported in the Tamale Metropolis, where N is the sample size, Z (statistic) = 1.96, p (compliance rate) = 0.262, and d (margin of error) = 0.05:

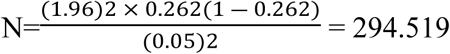

The calculated sample size was therefore 295. After adding a 10% non-response rate, the final sample size obtained was 324.

Simple random sampling was used to select the schools and the adolescent girls, while purposive sampling was used to select the school health coordinators and health staff. The lottery method was used to select the schools. The names of all junior high schools in the district were written on pieces of paper and placed in a bowl. With blind draw, the schools were randomly picked one by one without replacement until the expected number of 20 schools were reached. The final sample size of 324 was proportionally allocated to the 20 schools based on the enrolment of girls in each school. The study respondents were selected from each class through simple random sampling using random numbers generated for each class using the girls’ attendance register. Twenty-one school health coordinators in the selected schools and 14 health staff in charge of the school health services (6 oversaw two schools each) were purposively sampled to participate in the study. The proportional allocation of the sample size based on school enrolment led to the application of sampling weights during analysis to ensure that larger schools (with more girls) did not unduly influence results compared to smaller schools.

### Study Variables

The dependent variable was IFA compliance level (intake). The independent variables were knowledge on anaemia, knowledge on the IFAS program, socio-demographic factors of girls (e.g. age, marital status, level of education, and occupation of parents).

### Data Collection Tool and Technique

A structured pre-tested questionnaire was used to collect the data with the help of interviewer-assisted questionnaire administration. The data collection tool was adopted and modified from similar published studies [24, 25, 26,27]. Pretesting of the data collection tool was done in Nanumba North District, since it shares similar characteristics with the study area.

A prior permission was sought from headmasters and health coordinators of the selected schools through formal writing. The questionnaire, which was mostly closed-ended with multiple choice and dichotomous responses, was divided into sections and administered to both adolescent girls and teachers as well as health staff. The expected duration for participation in this study was 15 to 20 minutes per respondent, and the entire duration for the data collection and the study started on 1^st^ August 2022 to 28^th^ February 2023.

One-day training was organized for the data collectors on 26^th^ July 2022. They were trained on the objectives of the study, how to ensure confidentiality of the data, and how to fill the questionnaire. The data collectors were also trained on how to initiate the interview process by reading the informed consent to every respondent for voluntary participation. After data collection, the investigators reviewed each questionnaire to ensure the accuracy and completeness of the data collected.

### Data Analysis

The data was coded and analyzed using STATA 17.0 (Stata Corp, College Station, TX). Sociodemographic characteristics, level of compliance, and knowledge of anaemia and IFAS program was first presented in text, figures, and tables using descriptive statistics such as frequencies and percentages. Bivariate logistic and Multivariate logistic regression tests were performed to determine the association between the sociodemographic characteristics, knowledge on anaemia and IFAS program and the outcome variable (compliance level). Crude and adjusted odds ratios with their respective confidence intervals (95%) were computed to determine the strength of association of each variable.

IFAS compliance level was measured based on the number of tablets consumed in the past 7 weeks to the conduct of this study. Adolescent girls who consumed at least five tablets of the expected dose in the previous 7 weeks (1 tablet per week) which is equivalent to consuming 70% of the expected dose before the day of the data collection were considered compliant. Adolescent girls who consumed less than five tablets were considered noncompliant [28]. Records kept by the school health coordinators were used to validate each respondent’s compliance status.

A correct response was scored “1” for respondents who answered at least one cause (s) of anaemia, signs and symptoms of anaemia, consequences of anaemia, and prevention of anaemia correctly and an incorrect response was scored “0” on anaemia. Respondents who were able to get “1” in all the four questions were considered to have good level of knowledge. Knowledge of the IFAS program was scored “1” for good knowledge and an incorrect answer was scored “0” for poor knowledge item on why they are given the tablet, benefits of taking IFA tablet, food that inhibits iron and folic acid absorption, side effects of taking IFA tablets, and why girls are given the iron and folic acid tablet. Respondents who answered correctly at least four question (s) were those considered having good knowledge on the IFAS program while those who scored below the four marks were considered as having poor knowledge [29]. The study sought to identify and categorize levels of knowledge among respondents on both anaemia and IFA program, and acknowledging at least one correct answer allowed for the recognition of some baseline understanding. P-value less than 0.05 was used as cut-off point to see the presence of statistically significant association.

### Ethical Considerations

Ethical clearance for the study was sought from the Ghana Health Service Ethics Review Committee (Protocol number: **GHS-ERC:043/09/22**). Approval letter for the study was granted by the Nanumba South District Health Directorate of Ghana Health Service, while permission to go into the school was obtained from the Nanumba South District Education Office. Written informed assent was obtained from underage participants (those below 17 years) in their schools while another written informed consent was obtained from the parents of the girls. Selected girls were given the informed consent forms to send home for their parents to thumbprint/sign signaling their agreement for their child to participate in the study.

## Results

### Socio-demographic Characteristics of Respondents

From Table 1, the total number of respondents (adolescent girls) in this study was 324 with a mean age of 16.15 (±2.06). More than half (56.8%) of the respondents were within the age range of 10 to16 years with a little above two-third (68.2%) in form 2. Also, more than half (57.7%) of the respondents were from Kokomba ethnic group, while some 54.9% of them were affiliated to the Christian religion. Most (81.2%) of the respondents’ mothers had no formal education. In addition, less than three-quarters (71.0%) of the respondents’ fathers had no formal education. Finally, while a little above two-third (67.3%) of the mothers were farmers, majority (80.6%) of the fathers were engaged in the same occupation (farming).

**Table 1:** Socio-demographic characteristics of adolescent schoolgirls, Nanumba South District, Northern region, Ghana.

| Variables | Frequency (n=324) | Percentage (%) |
| --- | --- | --- |
| **Age (Years)** ($\bar{x}$: 16.15, SD: 2.06) | | |
| 10-16 | 184 | 56.8 |
| 17-25 | 140 | 43.2 |
| **Class** |  |  |
| JHS 1 | 103 | 31.8 |
| JHS 2 | 221 | 68.2 |
| **Ethnicity** |  |  |
| Dagomba | 119 | 36.7 |
| Fulano | 11 | 3.4 |
| Kokomba | 187 | 57.7 |
| Others | 7 | 2.2 |
| **Religion** |  |  |
| Christianity | 178 | 54.9 |
| Islam | 126 | 38.9 |
| Traditional | 20 | 6.2 |
| **Mother's level of education** |  |  |
| No formal education | 263 | 81.2 |
| Basic/Primary | 41 | 12.7 |
| Higher education | 20 | 6.1 |
| **Father's level of education** |  |  |
| No formal education | 230 | 71.0 |
| Basic/Primary | 40 | 12.4 |
| Higher education | 54 | 16.6 |
| **Mother's Occupation** |  |  |
| Farmer | 218 | 67.3 |
| Trader | 79 | 24.4 |
| Others | 27 | 8.3 |
| **Father's Occupation** |  |  |
| Farmer | 261 | 80.6 |
| Trader | 25 | 7.7 |
| Others | 38 | 11.7 |

### Level of knowledge on iron and folic acid supplementation among adolescent girls

From Table 2, the majority (59.0%) of the respondents indicated that they have heard about anaemia. Among those who have heard about anaemia, most of them heard it from their schoolteachers (52.4%), followed by those who heard it from health personnel (41.4%). The most common symptoms of anaemia identified in this study were pale skin (40.3%). In addition, the most major causes of anaemia among adolescent girls were identified to be iron deficiency, heavy menstrual bleeding, and infection (29.3%, 21.5%, and 13.1% respectively). Even though almost one-quarters (24.1%) of the respondents knew that anaemia decreases work capacity, and impact on school performance (23.6%) and delay mental and physical development (20.4%), some (31.9%) did not know any effect (s) of anaemia on adolescent girls. Moreover, more than one-quarter (28.3%) of the respondents knew that anaemia can be prevented by eating iron-rich foods. However, less than two-third (56.2%) of the respondents could not identify any iron-rich foods. Moreover, a little below two-third (68.5%) of the respondents did not also know the food that aid iron absorption, while less than three-quarter (70.4%) of them couldn’t also identify any beverage which decrease iron absorption. Finally, a greater proportion (96.3%) of the respondents indicated that they have heard about IFA supplementation.

**Table 2:** Knowledge of adolescent girls on anaemia and the IFAS program, Nanumba South District, Northern region, Ghana.

| Variables | Frequency (n=324) | Percentage (%) |
| --- | --- | --- |
| **Heard anaemia** |  |  |
| No | 133 | 41.0 |
| Yes | 191 | 59.0 |
| **Source of information\*\*** |  |  |
| Health personnel | 79 | 41.4 |
| Family members | 3 | 1.6 |
| Friends/neighbors | 5 | 2.5 |
| Media | 4 | 2.1 |
| School teacher | 100 | 52.4 |
| **Symptoms of anaemia\*\*** |  |  |
| More likely to become sick | 42 | 22.0 |
| Pale skin (pallor) | 77 | 40.3 |
| Weakness | 43 | 22.5 |
| Don't know | 29 | 15.2 |
| **Causes of anaemia\*\*** |  |  |
| Iron deficiency | 56 | 29.3 |
| Folic acid deficiency | 19 | 9.9 |
| Infection | 25 | 13.1 |
| Heavy menstrual bleeding | 41 | 21.5 |
| Don't know | 50 | 26.2 |
| **Effects of anaemia on adolescent girls\*\*** |  |  |
| Decreased work capacity | 46 | 24.1 |
| Delay of mental and physical development | 39 | 20.4 |
| Impact on learning (school performance) | 45 | 23.6 |
| Don't know | 61 | 31.9 |
| **Prevention of anaemia\*\*** |  |  |
| Eat iron-rich foods | 54 | 28.3 |
| Eat vitamin C-rich food during/after meals | 34 | 17.8 |
| Supplementation | 27 | 14.1 |
| Treat other causes of anaemia | 12 | 6.3 |
| Hygiene | 19 | 9.9 |
| Don't know | 45 | 23.6 |
| **Iron-rich foods** |  |  |
| Fish and seafood | 58 | 17.9 |
| Flesh meat | 38 | 11.7 |
| Organ meat | 26 | 8.0 |
| Organ meat Fish and seafood | 20 | 6.2 |
| Don't know | 182 | 56.2 |
| **Food helps iron absorption** |  |  |
| Vitamin C-rich foods | 102 | 31.5 |
| Don't know | 222 | 68.5 |
| **Beverage which decreases iron absorption** |  |  |
| Coffee | 59 | 18.2 |
| Tea | 37 | 11.4 |
| Don't know | 228 | 70.4 |
| **Heard about IFA supplementation** |  |  |
| No | 12 | 3.7 |
| Yes | 312 | 96.3 |

\*\* means the denominator used for these variables was less than the total sample used in the study.

In Fig 1, majority (32.8%) of the adolescent girls cited regulation of menstruation as the major benefit they have gotten from taking the IFA tablet. It was further found that about 20.6% thinks IFA tablets have the following benefits: regulate menstruation, improve their concentration and performance in class, increase appetite, feel stronger, reduce dizziness, and reduced fatigue among them, while 19.5% did not perceived any benefit (s) they have gotten from taking the IFA tablet.

**Fig 1:**
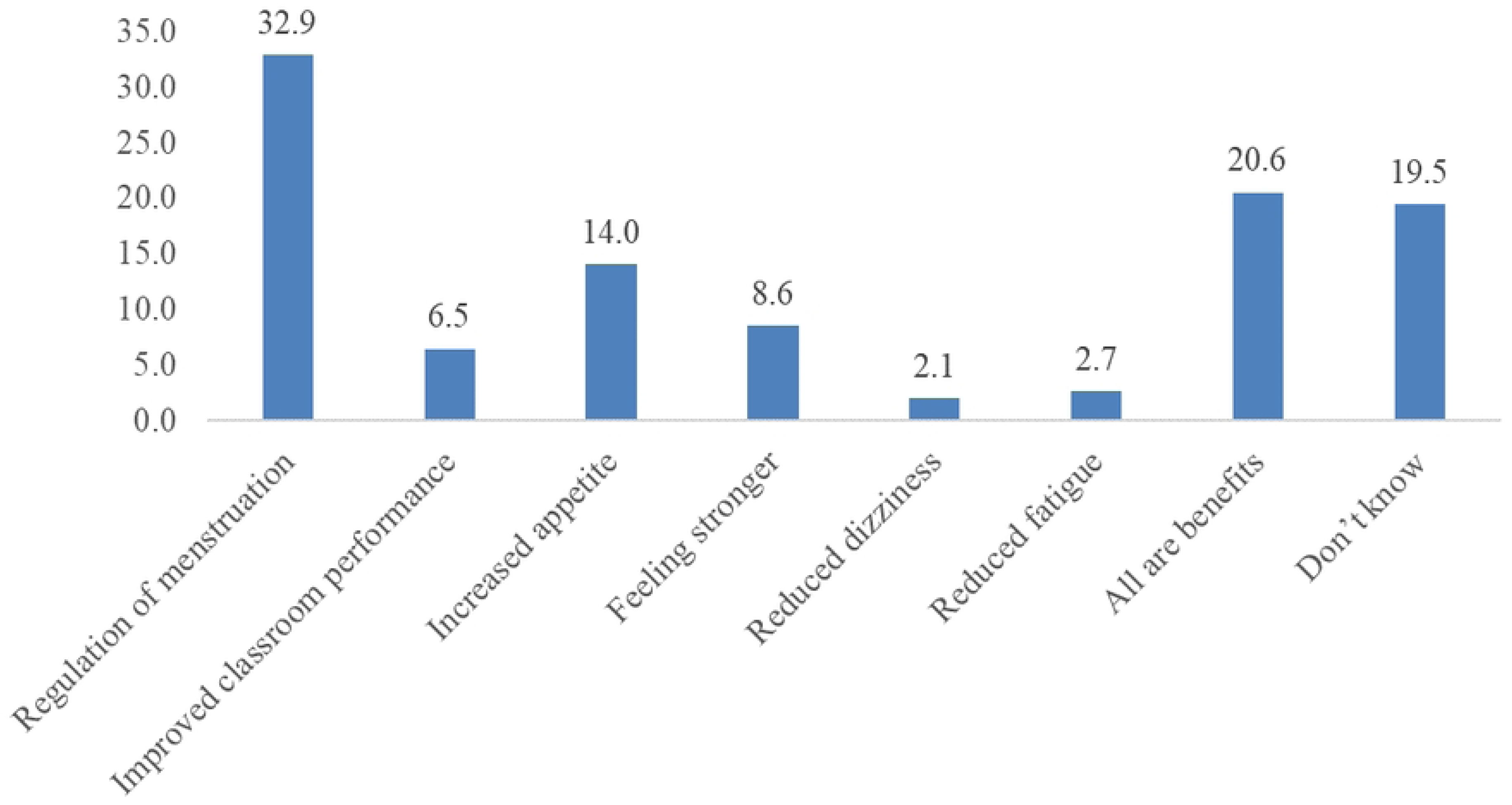
Benefits of consuming IFA tablets.

The overall level of knowledge of respondents on IFA supplementation and its benefits was generally found to be good (88.0%) (Fig 2).

**Fig 2:**
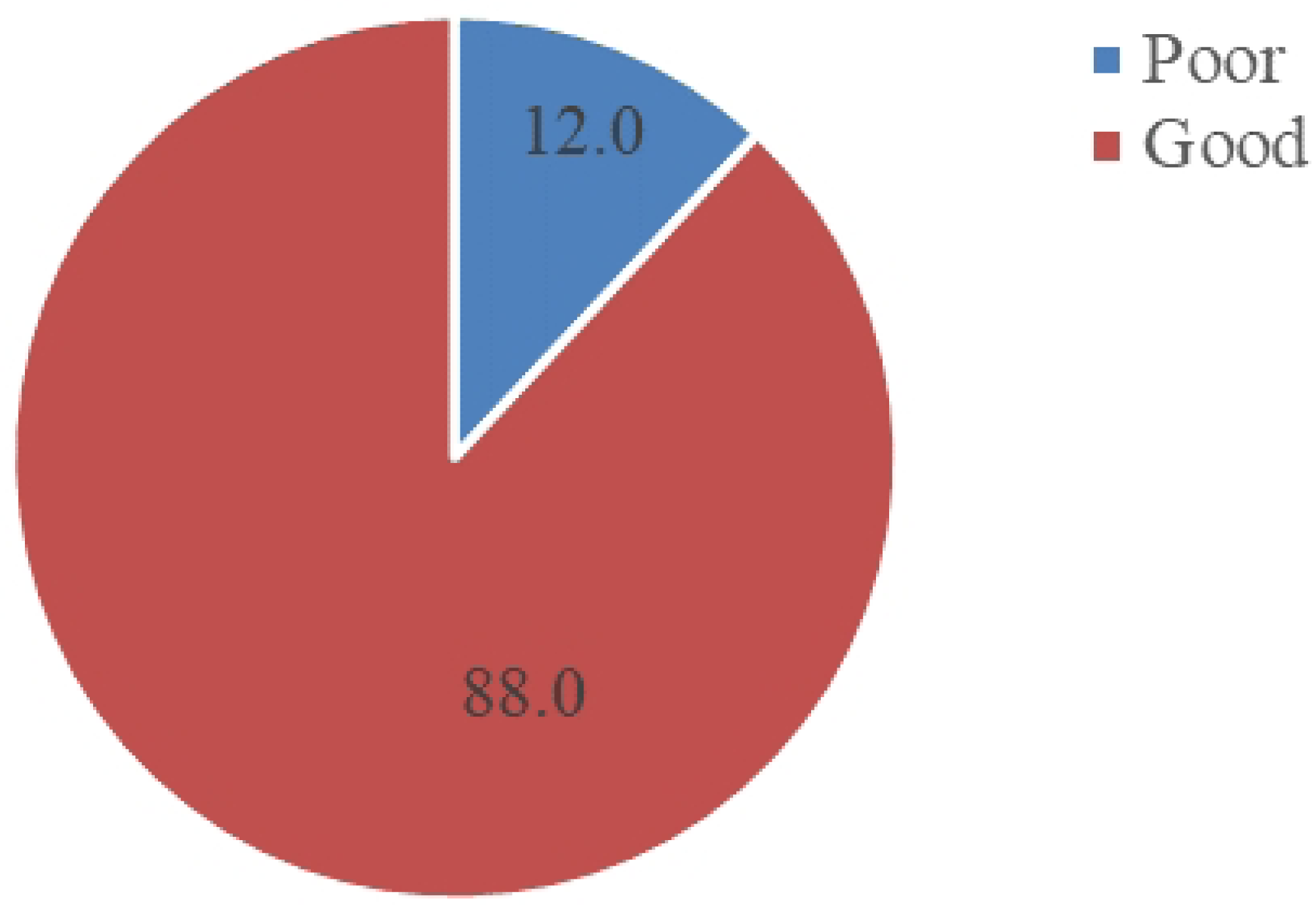
Level of Knowledge of in-schoolgirls about IFAS.

### Attitudes and Beliefs towards IFA tablets intake among adolescent girls and teachers

From Fig 3, most (85.2%) of the respondents said there were no side effects associated with the consumption of IFA tablet. However, some 4.6%, 3.7%, and 3.4% stated respectively that stomachache, black stool, and nausea were some of the side effects they experienced with the consumption of IFA tablet, while few (2.47%) cited headache.

**Fig 3:**
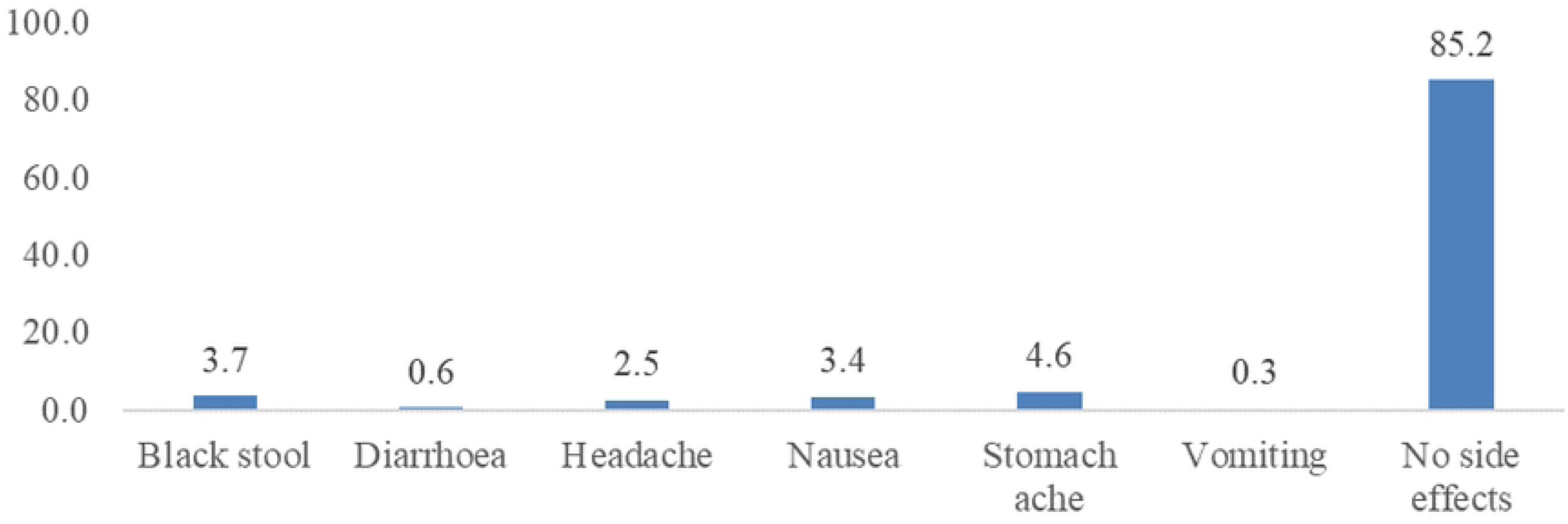
Perceived side effects of IFA supplement.

The results in Table 3 showed that a little above half (52.4%) of the respondents (teachers) always counsel their students on the IFA consumption, and many of them made the students consume their tablet in front of the teachers (90.5%). All the teachers have confirmed that the supply of IFA tablets to them was regular (100.0%). There was proper record keeping on the IFA supplements given to the students among majority of the teachers (85.7%) while many of the teachers were oriented on the IFA program (85.7%). In addition, most of the teachers noted that students did resist the consumption of IFA tablets (90.5%). Moreso, many of the respondents however indicated that the IFA program served as an additional work burden on them (90.5%). Finally, the majority (90.5%) of the teachers also said they gave the IFA tablets to students who were present on the day (s) the IFA tablets were given on the following day when they are present.

**Table 3:** Views of teachers on Attitude of and challenges towards the use of IFA supplementation, Nanumba South District, Northern region, Ghana.

| Variables | Frequency (n=21) | Percentage (%) |
| --- | --- | --- |
| Counselled on IFA consumption |  |  |
| Yes always | 11 | 52.4 |
| Yes sometimes | 10 | 47.6 |
| **Students consumed the tablet in front of teacher** |  |  |
| No | 2 | 9.5 |
| Yes | 19 | 90.5 |
| **Regular supply of tablets** |  |  |
| Yes | 21 | 100.0 |
| No | 0 | 0.0 |
| **Maintained records on IFA** |  |  |
| No | 3 | 14.3 |
| Yes | 18 | 85.7 |
| **Orientated on IFA program** |  |  |
| No | 3 | 14.3 |
| Yes | 18 | 85.7 |
| **Students resisted in taking IFA tablets** |  |  |
| No | 2 | 9.5 |
| Yes | 19 | 90.5 |
| **IFAS program puts extra burden on your workload** |  |  |
| Yes always | 19 | 90.5 |
| Yes sometimes | 2 | 9.5 |
| **Students given IFA when they absent from school** |  |  |
| Yes | 19 | 90.5 |
| No | 2 | 9.5 |

### Regular supportive supervision and monitoring on IFA supplementation to schools

From Table 4, all the respondents (health workers) indicated that they supervised IFA intake on weekly basis, 14 (100.0%). Most, 11 (78.6%) also said they did carried activities such as distribution, health education and promotion of IFA activities. Many of the respondents also said they regularly involved other departments, 9 (64.3%) in IFA activities. The reporting format of IFA activities were up to date among many of the health workers interviewed in this study, 11 (78.6%) while many of the respondents indicated they used IFA activities data in making decisions, 12 (85.7%).

**Table 4:**
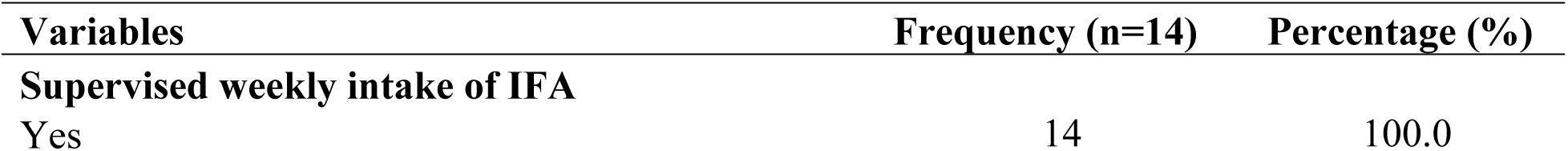
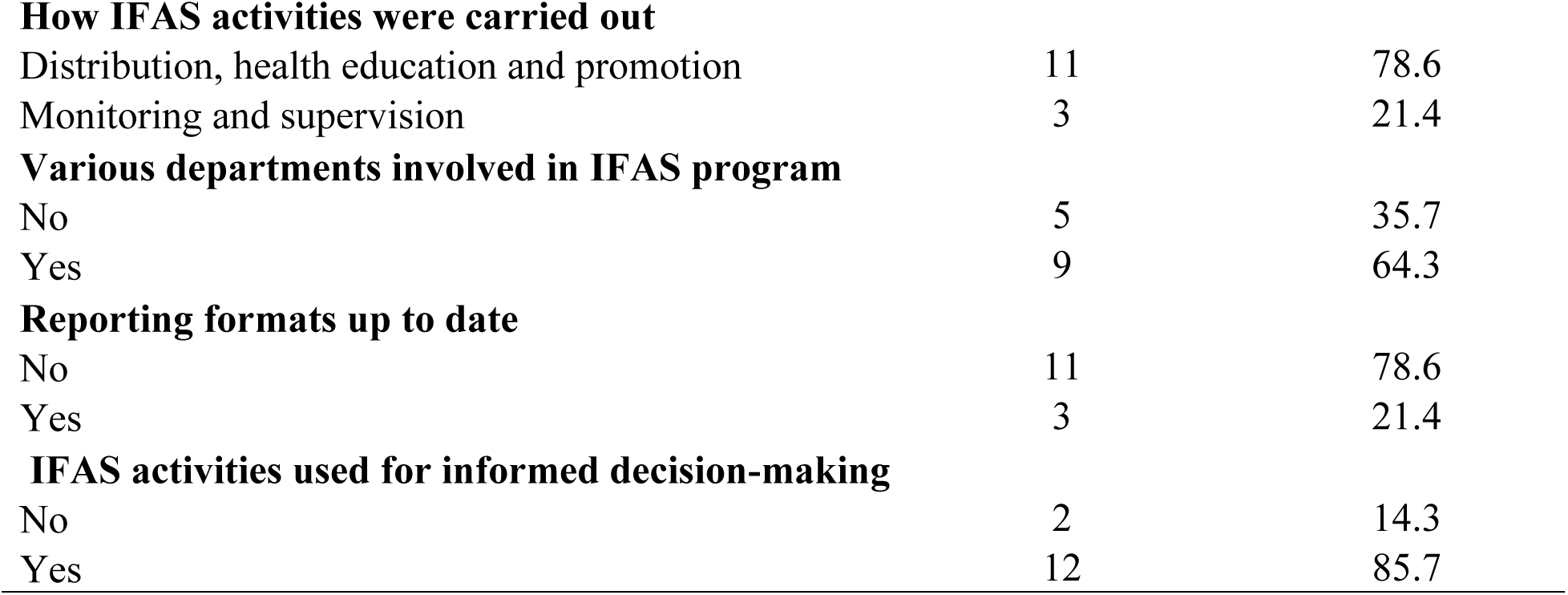
How regular health staff embark on IFA supplementation activities, Nanumba South District, Northern region, Ghana.

| Variables | Frequency (n=14) | Percentage (%) |
| --- | --- | --- |
| **Supervised weekly intake of IFA** |  |  |
| Yes | 14 | 100.0 |
| **How IFAS activities were carried out** |  |  |
| Distribution, health education and promotion | 11 | 78.6 |
| Monitoring and supervision | 3 | 21.4 |
| **Various departments involved in IFAS program** |  |  |
| No | 5 | 35.7 |
| Yes | 9 | 64.3 |
| **Reporting formats up to date** |  |  |
| No | 11 | 78.6 |
| Yes | 3 | 21.4 |
| **IFAS activities used for informed decision-making** |  |  |
| No | 2 | 14.3 |
| Yes | 12 | 85.7 |

### Acceptability (Compliance) of iron and folic acid supplements among adolescent girls

The results in Table 5 showed that overall, a greater proportion of the respondents complied with the IFAS consumption (65.7%), with most of the students taking the tablets under supervision (87.0%). The majority (95.4%) of the adolescent girls also took the IFA tablet on the first day it was given by their teachers. Furthermore, 86.1% of the adolescent girls noted that the last time they consumed the IFA tablet in school was 1-3 weeks ago before this survey. For adolescent girls who consumed the IFA tablets, the most common reasons why they took the tablets were because they: wanted to prevent anaemia (49.7%); received advice from their teachers, 108 (34.8%); and saw friends taking it (10.3%) respectively. Among those who did not take the IFA tablet in school (46.7%) cited the fact that they felt healthy. Most (55.6%) of the adolescent girls have never missed taking the IFA tablet in school. However, some 44.4% of the adolescent girls have missed taking the IFA tablet because they were absent from school (79.2%) or because they felt the tablet had a bad taste (9.7%).

**Table 5:** Compliance with IFAS among adolescent girls, Nanumba South District, Northern region, Ghana.

| Variables | Frequency (n=324) | Percentage (%) |
| --- | --- | --- |
| **Number of tablets taken in the past 7 weeks before the study** |  |  |
| <5 (Received some) | 111 | 34.3 |
| ≥5 | 213 | 65.7 |
| **Always take the tablet under supervision** |  |  |
| No | 42 | 13.0 |
| Yes | 282 | 87.0 |
| **Took IFA tablet on the first day** |  |  |
| No | 15 | 4.6 |
| Yes | 309 | 95.4 |
| **Last time taken IFA tablet** |  |  |
| 1-3 Weeks | 266 | 86.1 |
| 1 Month | 43 | 13.9 |
| **Reasons for taking IFA tablet** |  |  |
| Advice from teacher | 108 | 34.8 |
| It is free | 16 | 5.2 |
| Friends are taking it | 32 | 10.3 |
| It prevents anaemia | 153 | 49.7 |
| **Reasons for not taking the tablet** |  |  |
| Fear of side effects | 5 | 33.3 |
| My parents ask me not to take the tablet | 3 | 20.0 |
| I feels healthy | 7 | 46.7 |
| **Ever missed tablet in School** |  |  |
| No | 180 | 55.6 |
| Yes | 144 | 44.4 |
| **Reasons for missing** |  |  |
| Bad taste of tablet | 14 | 9.7 |
| Side effects | 11 | 7.6 |
| Was absent | 114 | 79.2 |
| Was not given IFA tablet | 5 | 3.5 |

From Table 6, adolescent girls whose fathers were farmers (85.5%), were of the majority group who complied with the IFA intake. The Pearson’s chi-square test established that father’s occupational level p=0.002) was the only variable which was significantly associated with adolescent girls’ IFA compliance.

**Table 6:** Factors associated with level of compliance of IFAS among adolescent girls, Nanumba South District, Northern region, Ghana.

| Variables | Compliance with IFAS |  | Total (N=324) | p-value |
| --- | --- | --- | --- | --- |
|  | Noncompliance | Compliance |  |  |
| Age (Years) |  |  |  |  |
| 10-16 | 58 (52.2) | 126 (59.1) | 184 (56.8) | 0.234 |
| 17-25 | 53 (47.8) | 87 (40.9) | 140 (43.2) |  |
| **Class** |  |  |  |  |
| JHS 1 | 38 (34.2) | 65 (30.5) | 103 (31.8) | 0.495 |
| JHS 2 | 73 (65.8) | 148 (69.5) | 221 (68.2) |  |
| **Ethnicity** |  |  |  |  |
| Dagomba | 37 (33.3) | 82 (38.5) | 119 (36.7) | 0.283 |
| Kokomba | 70 (63.1) | 117 (54.9) | 187 (57.7) |  |
| Others | 4 (3.6) | 14 (6.6) | 18 (5.6) |  |
| **Religion** |  |  |  |  |
| Christians | 59 (53 (2) | 119 (55.9) | 178 (54.9) | 0.813 |
| Islam | 44 (39.6) | 82 (38.5) | 126 (38.9) |  |
| Traditional | 8 (7.2) | 12 (5.6) | 20 (6.2) |  |
| **Mother's educational level** |  |  |  |  |
| No formal education | 89 (80.2) | 174 (81.7) | 263 (81.2) | 0.855 |
| Basic/Primary | 14 (12.6) | 27 (12.7) | 41 (12.7) |  |
| Others | 8 (7.2) | 12 (5.6) | 18 (6.1) |  |
| **Father's educational level** |  |  |  |  |
| No formal education | 79 (71 (2) | 151 (70.9) | 230 (71.0) | 0.331 |
| Basic/Primary | 17 (15.3) | 23 (10.8) | 40 (12.4) |  |
| Others | 15 (13.5) | 39 (18.3) | 54 (16.6) |  |
| **Mother's occupational level** |  |  |  |  |
| Farmer | 69 (62.2) | 149 (70.0) | 218 (67.3) | 0.347 |
| Trader | 32 (28.8) | 47 (22.1) | 79 (24.4) |  |
| Others | 10 (9.0) | 17 (8.0) | 27 (8.3) |  |
| **Father's occupational level** |  |  |  |  |
| Farmer | 79 (71.2) | 182 (85.5) | 261 (80.6) | **0.002** |
| Trader | 16 (14.4) | 9 (4.2) | 25 (7.7) |  |
| Others | 16 (14.4) | 22 (10.3) | 38 (11.7) |  |
| **Awareness of anaemia** |  |  |  |  |
| Yes | 58 (52.3) | 133 (62.4) | 191 (58.9) | 0.077 |
| No | 53 (47.8) | 80 (37.6) | 133 (41.1) |  |
| **Knowledge on anaemia** |  |  |  |  |
| Good | 55 (94.8) | 114 (85.1) | 169 (88.0) | 0.056 |
| Poor | 3 (5.2) | 20 (14.9) | 23 (12.0) |  |
| **Knowledge on IFAS program** |  |  |  |  |
| Good | 107 (96.4) | 208 (97.7) | 315 (97.2) | 0.514 |
| Poor | 4 (3.6) | 5 (2.4) | 9 (2.8) |  |

From Table 7, the odd of compliance with IFAS among respondents who among adolescent girls 17-25 years was 56% times lower compared to those who were 10-16 years (AOR = 0.44 (95% CI: 0.21-0.96) p = 0.038). Also, Kokomba girls were 83% less likely to comply to IFA consumption as compared to Dagomba adolescent girls (AOR = 0.17 (95% CI: 0.03-0.89) p = 0.036).

**Table 7:**
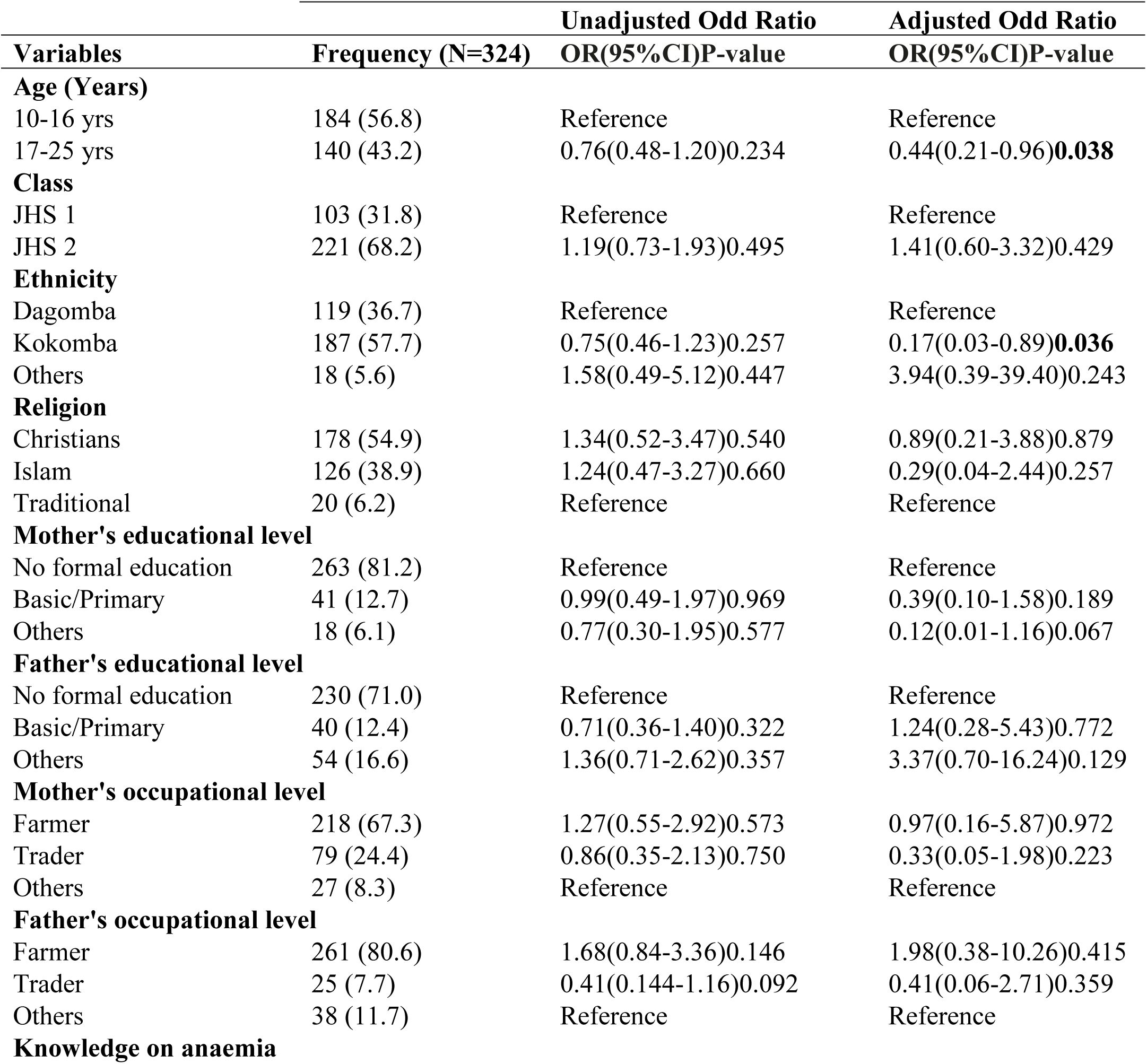
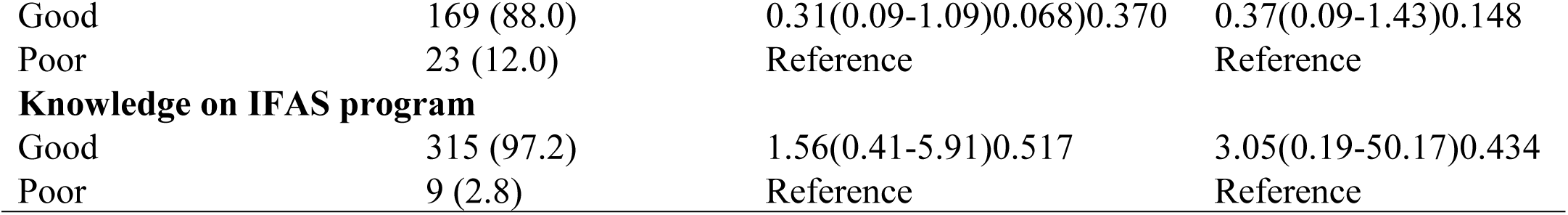
Predictors of level of compliance of IFAS among adolescent girls, Nanumba South District, Northern region, Ghana.

## Discussion

### Level of knowledge on the benefits of iron and folic acid supplementation among adolescent girls

In this present study, it was found that the level of knowledge of respondents on IFA supplementation and its benefits was generally good (88.0%). This finding contradicted with what was reported in another study in Tamale, Ghana, that about 64.9% of adolescent girls had poor level of knowledge IFA supplementation [16]. However, among different population of pregnant women in Northwest Ethiopia, a lower figure of about 57.5% was reported to have good level of knowledge about anaemia [29].

We also found that less than two-fifths (32.9%) of the adolescent girls noted that the major benefit they have gotten from taking the IFA tablet was regulation of menstruation only. It was further found that 20.55% of the girls knew that regulation of menstruation, improving concentration and performance in class, increasing appetite, feeling stronger, reducing dizziness, and reducing fatigue were all benefits of taking the IFA tablets. Similarly to these findings, it was revealed in the United States that reduced fatigue, increased appetite, and improved concentration were respectively the major cited benefits of IFA tablets [30]. However, some 19.5% did not know any benefit (s) they have gotten from taking the IFA tablet.

### Attitudes and Beliefs towards IFA tablets intake among adolescent girls and teachers

In this study, most (85.2%) of the respondent girls indicated that the consumption of IFA tablet had no associated side effects. This differed from the report from a cross-sectional study in Ethiopia which found that one of the main reasons for non-adherence to iron folate supplementation was fear of side effect [29]. However, some respondents in this current study mentioned stomachache, black stool, nausea, and headache as some of the perceived side effects they experienced with the consumption of the IFA tablet. Consequently, these side effects could threaten the success of the IFAS program. A similar finding was reported in India and US that the major negative factors for the program were fear of adverse effects such as nausea and vomiting, attribution of abdominal pain due to other causes of IFA tablets [27,30]. This present study’s finding also aligned in parts with what was reported in Iran, that an upset feeling, vomiting, abdominal discomfort, unpleasant taste of the pills, and feeling of drowsiness/dizziness due to the poor quality of IFA tablets were the other main reasons for refusing the pills [31].

Moreover, in this present study, a little above half (52.4%) of the respondent teachers did counsel their students to take the IFA tablet. It was also found that a vast majority (90.5%) of the teachers made their students consumed the IFA tablets in front of them. This is encouraging as less teachers did not follow the IFAS program recommendation that teachers should directly supervise the adolescent girls to swallow the IFA tablet [32]. In schools, supervising students to ingest the IFA tablet is considered one of the most important aspects of the IFAS program [33]. Without supervision, the possibility of school children throwing the tablet away is reportedly likely to be very high [34]. The supply of IFA tablets to the schools was reported to be regular as found among all the respondent teachers. This study also found that proper adherence to IFA records keeping was observed among majority of the teachers (85.7%), while many of the teachers were oriented on the IFA program (85.7%). This finding is important because it is most likely to give an opportunity to build the teachers’ capacity to educate students to consume IFAS.

It was further revealed in this study that most of the adolescent girls initially resisted the consumption of IFA tablets (90.5%). This is contrary to what was reported in a previous study that there was no resistance in taking tablets from students [24]. Moreover, many of the respondent teachers complained that the IFA program was an additional work burden on them (90.5%). This finding agreed with what was reported in India that IFAS program was an extra workload on teachers [27]. To ensure that the student girls did not miss the tablets, the majority (90.5%) of the teachers gave the IFA tablets to those who absent themselves from school on the day (s) the IFA tablets were given, when they come to school the next day.

### Supportive supervision and monitoring on IFA supplementation to schools

In this current study, it was revealed that all the respondents (health workers) supervised IFA intake on weekly basis. Most of the respondents carried out activities such as distribution, health education and promotion of IFA in the various schools. Many of the teachers were found to always regularly involved other departments in IFA activities. This current study finding did not support what was reported in India that lack of coordination and ownership among the various departments were noticed [35]. The reporting format of IFA activities were also found to be up to date among many of the health workers interviewed in this study. This contradicted with the finding in a previous study that some of the reporting formats were outdated and missed section for IFA syrup [35]. It was further revealed that IFA activities data obtained from the various schools in this study were usually used in making informed decisions towards the successful implementation of the IFA program as opposed to what was previously reported that focus had been on the collection of data, but its utilization for informed decision-making and policy decisions was lacking [35].

### Compliance with iron and folic acid supplements with IFAS among adolescent girls

In this study, almost two-third (65.7%) of the respondents complied with the IFAS intake, which was quite recommendable. However, this compliance was lower as compared to similar a study which showed 81.7% among adolescent girls in Karaga District, Ghana [36]. This finding also differed from what was reported among high school girls that adherence to IFAS was only 37.2% [34]. High compliance in this study, which is expected to help improve iron status among the student girls could be a result of stock availability of the IFA tablet as noted in the schools.

Furthermore, the majority (95.4%) of the adolescent girls took the IFA tablet on the first day it was given by their teachers. Many (86.1%) of the girls however noted that the last time they have consumed the IFA tablet in school was 1-3 weeks ago before this study. The most common motivating factors for consuming the IFA tablet by the girls were because they wanted to prevent anaemia and that they were being advised by their teachers respectively. The adolescent girls who did not take the IFA in school said it was because they perceived they were healthy (46.7%). This present finding confirmed what was previously reported in India that one of the main reasons that led to their respondents not consuming iron and folic acid were the feeling that they were healthy [24].

Even though more than half (55.6%) of the girls never missed taking the IFA tablet in school, it was found that a significant proportion of 44.4% of them missed taking the IFA tablet because they were absent from school, or they felt the tablet had a bad taste. A similar finding was reported in Indonesia that motivation to take IFA tablet declined when respondents perceived the odour of tablet was unpleasant and experienced side effects [37].

This study also found that father’s occupational level was the only variable which had a statistically significance association with adolescent girls’ level of IFA compliance. These findings did not agree with what was reported by Dubik and colleagues who noted that the level of education and occupation of mothers of adolescent girls, awareness on anaemia, and good knowledge of anaemia and of the IFAS program were significantly associated with IFAS compliance [16]. The odds of compliance with the IFAS among respondents who were 17-25 years were 56% times lower compared to those who were 10-16 years in this study. This could be because adolescent girls who were older were curious about their health and tried taking the necessary measures to prevent themselves from sickness. In addition, it was revealed that respondents who were Dagomba adolescent girls had a higher likelihood of compliance to IFAS as compared to Kokomba who were 83% less likely to comply to IFA consumption. This present study findings did not also corroborate with what was reported in the Karaga District of Northern Region that apart from socio-economic status of respondents, guardian’s level of education and in or out of school adolescent, all other variables did not have any statistically significant relationship with the adherence to the required number of Iron and Folic Acid intake per month [36].

## Conclusion

In conclusion, we found that most of the respondents complied with the IFAS (65.7%). Also, while most respondents reported no associated side effects with IFA tablet consumption, a good proportion perceived side effect such as stomachache, black stool, nausea, and headache. The study further underscored the role of teachers in counseling and monitoring students, with a significant portion ensuring that students consume IFA tablets in their presence.

Interestingly, a considerable number of students initially resisted IFA tablet consumption, but a majority eventually complied because of a series of encouragement from their teachers. The demographic variable of father’s occupational level emerged as the sole factor significantly associated with adolescent girls’ level of IFA compliance. The odds of compliance were notably lower among girls aged 17-25 compared to those aged 10-16 years and Kokombas had lower odds of IFA compliance compared to the Dagombas.

Health workers played a crucial role in IFA program implementation, regularly supervising intake and maintaining up-to-date reporting formats. This study encompassed not only the students’ experiences and compliance with IFAS, but also the pivotal roles of teachers and health workers in ensuring its success were notified in this study.

### Recommendations

1. Efforts should be made by all stakeholders to involve and encourage local community leaders and religious leaders in IFAs implementation. This will help to motivate the parents and students to take IFA regularly and to address the benefits of IFA.
2. Future research should be conducted to determine the relationship between IFAs intake and level of anaemia status among adolescent girls.

## Data Availability

The data for this study is available and will be made accessible upon reasonable request from the corresponding via.

## Acknowledgement

We acknowledge the District Director of Education, Nanumba South District, and all headteachers and class teachers at the schools, for respectfully accepting us to conduct this study in their schools. Finally, we wish to thank our research participants (students, teachers, and health workers) for their cooperation and willingness to take part in the study. We finally acknowledge the good work done by our data collection assistants for helping us administer the questionnaires successfully.

## Limitations of the Study

Most of the results in this study were self-reported by adolescent girls. There was a possible tendency to recall bias and exaggeration, hence, the result was interpreted with caution. The study did not also measure Hb levels of the adolescent girls to determine the impact of the IFAS on their Hb levels. Finally, this study could not include the schoolteachers and local health workers’ knowledge and attitude towards this program and suggestions for improvement.

